# From Pre-test and Post-test Probabilities to Medical Decision Making

**DOI:** 10.1101/2024.02.14.24302820

**Authors:** Michelle Pistner Nixon, Farhani Momotaz, Claire Smith, Jeffrey S. Smith, Mark Sendak, Christopher Polage, Justin D. Silverman

## Abstract

**Background:** A central goal of modern evidence-based medicine is the development of simple and easy to use tools that help clinicians integrate quantitative information into medical decision-making. The Bayesian Pre-test/Post-test Probability (BPP) framework is arguably the most well known of such tools and provides a formal approach to quantify diagnostic uncertainty given the result of a medical test or the presence of a clinical sign. Yet, clinical decision-making goes beyond quantifying diagnostic uncertainty and requires that that uncertainty be balanced against the various costs and benefits associated with each possible decision. Despite increasing attention in recent years, simple and flexible approaches to quantitative clinical decision-making have remained elusive.

**Methods:** We extend the BPP framework using concepts of Bayesian Decision Theory. By integrating cost, we can expand the BPP framework to allow for clinical decision-making.

**Results:** We develop a simple quantitative framework for binary clinical decisions (e.g., action/inaction, treat/no-treat, test/no-test). Let ***p*** be the pre-test or post-test probability that a patient has disease. We show that ***r\** = (1 *− p*)*/p*** represents a critical value called a decision boundary. In terms of the relative cost of under- to over-acting, ***r\**** represents the critical value at which action and inaction are equally optimal. We demonstrate how this decision boundary can be used at the bedside through case studies and as a research tool through a reanalysis of a recent study which found widespread misestimation of pre-test and post-test probabilities among clinicians.

**Conclusions:** Our approach is so simple that it should be thought of as a core, yet previously overlooked, part of the BPP framework. Unlike prior approaches to quantitative clinical decision-making, our approach requires little more than a hand-held calculator, is applicable in almost any setting where the BPP framework can be used, and excels in situations where the costs and benefits associated with a particular decision are patient-specific and difficult to quantify.

## 1 Introduction

The Bayesian Pre-test/Post-test Probability (BPP) framework provides a quantitative framework for expressing diagnostic uncertainty and updating that uncertainty given clinical signs or test results. The BPP framework is used as a tool at the bedside, in the classroom, and as part of medical research [1]. Yet the BPP framework is only one part of what could be a much larger toolkit that clinicians, educators, and researchers could employ.

Though often discussed in the context of clinical decision-making [2, 3], the BPP framework is insufficient for it: while pre- and post-test probabilities represent beliefs in the disease state of a patient, clinical decision-making requires that diagnostic uncertainty be balanced against various cost and benefits factors. For example, a decision to treat must consider not only the probability that a patient has the disease in question but also the safety and efficacy of the proposed treatment. Yet these costs and benefits are often difficult to quantify, e.g., consider quantifying the psycho-social costs of a prophylactic bilateral mastectomy in a BRCA1 carrier. In short, quantitative approaches to clinical decision-making goes beyond the BPP framework and requires balancing diagnostic uncertainty with patient-specific, difficult-to-quantify cost and benefit factors.

Despite the challenges, there is a rich history of methods that have been developed for quantitative clinical decision-making. Yet most of these methods have either required complex computational models that cannot be easily performed at the bedside [4–6] or are specialized to a particular decision task (such as aspirin treatment for pre-eclampsia prevention [7]). The key exception is the almost 50 year old work by Pauker and Kassirer [8] which provides a flexible general purpose approach to action/inaction (e.g., treat/no-treat or test/no-test) decisions that is simple enough to be applied at the bedside or discussed easily in a classroom. The Pauker and Kassirer (PK) framework combines diagnostic uncertainty with quantified costs and benefits to determine a treatment threshold based on probability: a decision boundary between action and inaction stated in terms of diagnostic uncertainty. Despite the maturity of that work, it has not seen widespread adoption. While it is still discussed in certain educational settings (e.g., Newman and Kohn [9], Chapter 2), when compared to the BPP framework, it is largely unknown. We argue that the major limitation of the PK framework, which has limited its adoption, is its requirement that all costs and benefits be pre-specified and explicitly quantified.

The purpose of this article is to review the PK framework and to propose a simple reformulation that addresses its major limitation. In brief, the PK framework requires that all costs and benefits be specified *a priori*. With these specified costs, a decision boundary *p*^*\**^ can be calculated which represents the probability of disease at which action and inaction are equally optimal. A clinician-calculated probability *p* can then be compared to *p*^*\**^ to determine whether action or inaction are warranted. Here we show that the decision boundary *p*^*\**^ can be reformulated in such a way that it can be calculated without requiring pre-specification of all costs and benefits. We call this the *Simplified PK* (SPK) framework. We discuss the SPK framework in clinical and educational contexts through a series of hypothetical case studies. In addition, we illustrate its use as a research tool through a reanalysis of a study which found widespread overestimation of disease probabilities among clinicians [10]. Unlike prior analyses of that data, our results suggest that this over inflation may not be due to errors in clinical perception but instead a bluntness of the survey instrument used.

## 2 Methods

### 2.1 Probabilities and Odds

We use *p* to denote the probability that a patient is in an *actionable state*. The precise definition of actionable state will depend on the clinical decision under question. For example, when deciding whether or not to treat with an antibiotic, *p* could represent the probability that a patient has an infection treatable with the antibiotic. At times, it will be more natural to state our results in terms of odds rather than probabilities. Probabilities (*p*) can be calculated from odds (*o*) and vice versa using the following two relationships:

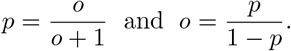

### 2.2 Review of the BPP Framework

Bayesian statistics provides a quantitative tool for updating prior beliefs (quantified as probabilities or odds) based on observed data. Compared to the full framework of Bayesian statistics, the BPP framework includes a critical simplification: the patient state is binary (e.g., disease or health as opposed to mild/moderate/severe disease). Under this simplification, Bayesian statistics reduces to the BPP framework.

Let *o*_pre_ (or *o*_post_) denote the odds of an actionable state before (or after) observing a particular diagnostic test or clinical sign. With this notation, the BPP framework can be written as

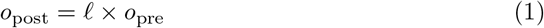

where *𝓁* is the likelihood ratio of the test or sign (a quantity capturing the evidence provided by the observed data). In short, given a clinician specified pre-test odds and given the likelihood ratio of a particular sign or test, the BPP framework computes an updated (post-test) odds. More discussion on the BPP framework in the context of medical decision-making can be found in Mark and Wong [11] or Armstrong and Metlay [12].

A note on notation: Much of what follows can be applied equally well to pre-test probabilities (or pre-test odds) as to post-test probabilities (or post-test odds). Therefore, we will often drop the subscript *pre* or *post* and simply write *p* or *o*.

### 2.3 BDT and the Pauker-Kassirer (PK) Framework

Bayesian Decision Theory (BDT) extends Bayesian statistics and provides a principled approach to making decisions under uncertainty. Compared to Bayesian statistics, BDT requires an additional user input: a cost function which takes two inputs and outputs a number between negative and positive infinity. The two inputs are the patient state and a potential action. The numerical output represents the *cost* or *loss* incurred by a given action and state of patient combination. Costs are a general concept and may incorporate medical, monetary, psycho-social, or even opportunity costs that may result from a given action. Negative costs are often called benefits. Were the patient state known exactly, BDT would reduce to finding the action that minimizes cost. Uncertainty in the state of the patient complicates the problem. To address this, BDT defines an optimal action (the “Bayes action”) as the action that minimizes the *expected cost* : the cost weighted by the probability of disease or no disease.

Optimizing expected cost often involves advanced numerical techniques, as seen in many prior articles in medicine [4–6]. We follow Pauker and Kassirer [8] and take a simpler approach. Beyond the standard simplifying assumption of the BPP framework (that the patient state is binary), we also assume that the considered action is binary (e.g., test/no test or treat/no treat). This implies that there are four combinations of action and patient state, each with an associated cost: accurate action (*c*_accurate action_, e.g., treating with disease), accurate inaction (*c*_accurate inaction_, e.g., treating with no disease), inaccurate action (*c*_inaccurate action_, e.g., treating with no disease), and inaccurate inaction (*c*_inaccurate inaction_, e.g., not treating with disease). Using shorthand, we denote these as *c*_*aa*_, *c*_*ai*_, *c*_*ia*_, and *c*_*ii*_, respectively. Letting that *p* denote the probability of an actionable state (e.g., disease), the expected cost of action (or inaction) decisions is then given by:

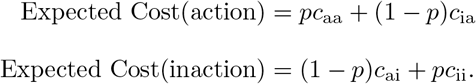

The Bayes action is action or inaction depending on whether Expected Cost(action) or Expected Cost(inaction) is less.

Given the four cost values *{c*_aa_, *c*_ia_, *c*_ai_, *c*_ii_*}*, Pauker and Kassirer [8] suggest calculating the threshold probability *p*^*\**^ needed to warrant action. That is, *p*^*\**^ is the specific probability at which Expected Cost(action) and Expected Cost(inaction) are equal. A physician calculated probability *p* (e.g., calculated using the BPP framework) can be compared to *p*^*\**^ to determine if action or inaction is optimal. They showed that this threshold was equal to:

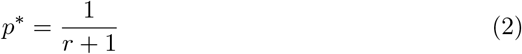

where

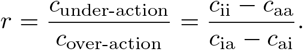

The term *c*_under-action_ represents difference in costs between inaccurate and accurate action when a patient is in a state that warrants action. Similarly, the term *c*_over-action_ represents the difference in costs between inaccurate and accurate action when the patient is in an state that warrants no action. In essence, these two terms represents the additional cost of choosing the wrong action over the correct. We refer to these two terms as the cost of under-action and over-action, respectively.

Overall, what we call the PK framework consists of first quantifying the four cost values, calculating the ratio of the costs of under- to over-action and then comparing the resulting action threshold *p*^*\**^ against a physician-calculated (or specified) probability of an actionable state *p*.

### 2.4 The SPK Framework

The major limitation of the PK framework is the need to pre-specify the four cost values in order to calculate the decision boundary *p*^*\**^. Especially when the costs or benefits of a potential action are patient-specific or even qualitative, it can be impossible to use the PK framework. Yet, we can mitigate this limitation of the PK framework by framing the same decision boundary in terms of a critical value for the ratio *r* = *c*_under-action_*/c*_over-action_ rather than a critical value for the probability *p*:

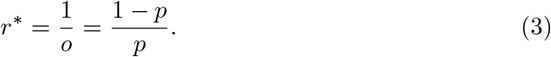

Equation (3) (SPK Framework) represents the same decision boundary as Equation (2) (PK Framework) but is presented differently (see Supplementary Section 1, Additional File 1 for derivation). Yet, we expect Equation (3) will be substantially easier to use than Equation (2). Consider than Equation (2) requires that all four costs values be specified in order to calculate *p*^*\**^. In contrast, Equation (3) requires only that a probability *p* or odds *o* be specified. Given the ubiquity of the BPP framework, we expect the later will be substantially easier than the former: any clinician already using the BPP framework can essentially calculate *r*^*\**^ for free (without further input).

There are two main ways to use Equation (3) depending on whether or not the four cost values *{c*_*aa*_, *c*_*ia*_, *c*_*ai*_, *c*_*ii*_*}* can be quantified. If those cost values cannot be specified, e.g., if the costs are fundamentally qualitative and patient-specific, then Equation (3) can be used as a qualitative tool to help incorporate diagnostic uncertainty into the decision-making process. As an example, consider a patient with a 20% probability of being in an actionable state. This implies a decision boundary *r\** = 4. In words, regardless of the particular action in question, *the cost of under-treating must be at least 4 times higher than the cost of over-treating in order to warrant action in this patient*. Even if the actual cost of under- and over-treating cannot be calculated, we expect this result may help frame the decision and catalyze discussions about costs and benefits. When costs can be quantified, clinicians can calculate *r* and compare it to *r\** to determine whether action or inaction is warranted. In Supplementary Section 1 (Additional File 1), we discuss how to compare *r* and *r\** in more detail.

## 3 Applications

We illustrate the SPK framework through a hypothetical case study of asymptomatic bacteriuria and a reanalysis of a recent study which found widespread misestimation of pre- and post-test probabilities by clinicians [10]. In Supplementary Section 2 (Additional File 1), we provide an additional case study designed to illustrate how subjective and objective cost factors can be combined within the SPK framework. All three of these applications are designed to highlight the use of the SPK framework in situations where the PK framework cannot be applied: where it is difficult to specify the four cost values *a priori*.

### 3.1 Case Study: Antibiotics for Asymptomatic Bacteriuria

Consider a healthy, non-pregnant, pre-menopausal 35-year-old woman diagnosed with asymptomatic gram negative bacteriuria by urine culture, with no recent history of Urinary Tract Infections (UTIs). Based on this presentation, prior work suggests that this patient has an approximately 6% probability of progressing from asymptomatic bacteriuria to symptomatic bacteriuria [13, 14]. Let this probability denote a post-test probability, where the state in question is whether the patient’s pathology will progress to symptomatic bacteriuria. We must decide whether to treat this patient with antibiotics at this time or wait and reevaluate if symptoms present.

Regardless of whether this 6% represents a pre-test or post-test probability, it is enough to calculate the decision boundary: 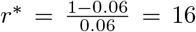. In words: *Given our diagnostic uncertainty, the cost of under-treating must be at least 16 times greater than the cost of over-treating to warrant treatment*.

In this situation, we expect that many clinicians will find it difficult to apply the PK framework directly: it is difficult to quantify the cost of under- and over-treating as there are many considerations including morbidity, monetary, and public-health (e.g., antibiotic resistance) costs. Even if all these costs could be specified exactly, combining them into an overall cost of over- and under-treating is not straightforward. Despite these challenges, we show that the decision boundary, particularly its translation into words, provides insights for contextualizing the decision in terms of diagnostic uncertainty. As a starting point, we suggest considering each type of cost individually (e.g., monetary, morbidity, and public health), from most to least important. If these costs agree on which decision is optimal, the choice of treatment decision is obvious. If they do not agree, we suggest careful consideration of which costs are most important and how strongly each cost supports its preferred decision.

In this case, we consider morbidity, public health, and monetary costs in that order. Either individually, or in concert with the patient, we would consider the following question: “Is the morbidity associated with under-treating at least 16 times greater than that of over-treating?” In our reading of the literature [15, 16], we expect that the morbidity associated with under-treating is likely higher than the morbidity associated with over-treating, yet we expect it is not 16 times greater especially when the low probability of progression to pyelonephritis and the probability of adverse reactions to antibiotics are considered. Therefore, considering morbidity alone we consider that no-treatment is warranted at this time. Additionally, after asking analogous questions about the monetary and public health costs, we believe the costs are higher in the case of over-treating. In sum, our suggestion would be to not treat this patient at this time but to pursue watchful waiting. This conclusion is supported by current treatment recommendations for asymptomatic bacteriuria in healthy non-pregnant persons [17].

### 3.2 Reevaluating the Source of Inflated Probability Estimates among Clinicians

In a study of 553 medical practitioners presented with four different clinical scenarios, Morgan et al. [10] found widespread over-estimation of both pre-test and post-test probabilities compared to objective probability estimates. The results of this study are significant and led to a conversation about potential biases affecting the observed inflation (for example, see Chaitoff [18]; Patel and Goodman [19]). Current hypotheses include various factors that confound physicians’ perceptions and interpretations of medical tests [10, 20]. For example, Kellner [20] suggests this over-estimation comes from less-experienced physicians included in the study cohort. While we do not doubt that such factors are at play, we hypothesize that the observed inflation may result from a bluntness of the survey instrument used to elicit physician probability estimates. What if the physicians surveyed included in their reported probabilities consideration of various cost factors: e.g., a physician asked to quantify the probability of disease might modify their probability estimates based the risk of missing a diagnosis. Assuming that the cost of under-treatment is greater than the cost of over-treatment, we expect that such conflation would lead reported probability estimates to exceed objective probability estimates, exactly as reported in Morgan et al. [10].

The SPK framework can be used to gauge the plausibility of this hypothesis. Let us suppose that our hypothesis is true: the Morgan et al. [10] survey elicited a physician’s *probability to act* (*p*_act_) rather than their estimation of the probability of disease (*p*_disease_). Further, let us assume that clinicians’ perceptions about disease probabilities are accurate: physicians’ true estimates of *p*_disease_ are equal to their objective values. Let us assume that the probability that a physician will act is proportional to their perceptions about the expected cost of action. Using this later assumption, we can calculate the physicians perceptions about the relative cost of under- to over-treating (the *implied cost-ratio*) as:

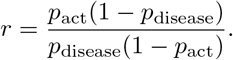

See Supplementary Section 3.1, Additional File 1 for derivation.

Below, we reanalyze the results of Morgan et al. [10] and calculate implied cost-ratios for three diseases studied in that work. We argue that the implied cost ratios are defensible given current evidence. In short, we are able to provide a model for the inflated probability estimates observed in Morgan et al. [10] based only on a bluntness of the survey instrument and without requiring overt errors in clinical perception.

#### 3.2.1 Re-analyzing Morgan et al

We calculated implied cost ratios for three of the four clinical scenarios studied in Morgan et al. [10]: pneumonia, breast cancer, and coronary artery disease. We excluded the asymptomatic bacteriuria scenario from our reanalysis due to concerns about the accuracy of the survey data (see Supplementary Section 3.2, Additional File 1 for details). The resulting cost-ratios are 15.8 for the pneumonia scenario, 16.7 for the breast-cancer scenario, and 21.4 for the coronary artery disease scenario.

We argue that these implied cost ratios could represent realistic clinical attitudes towards risk. To justify our opinion, we provide an example clinical argument justifying the 16.7 breast-cancer cost-ratio in Supplementary Section 3.3, Additional File 1. Moreover, we can compare to prior literature which studied physicians’ attitudes towards the risk of over- and under-treating patients with pneumonia [21]. Based on that prior literature, we estimate a pneumonia cost-ratio of 14.7 (see Supplementary Section 3.4, Additional File 1). We suggest that the correspondence between that result and the estimated results from Morgan et al. [10] is a strong argument that these implied cost ratios reflect potentially realistic clinician attitudes towards risk. Together, this suggests that the inflated probability reported by Morgan et al. [10] may not be due to altered physician perceptions but may instead be due to a bluntness of the survey instrument. Overall, our results strongly suggest that future research should consider that physicians may conflate probabilities of disease with factors affecting decisions to act.

## 4 Discussion

We introduced the Simplified PK (SPK) Framework as a flexible and easy-to-use tool for medical decision-making when there is uncertainty in the state of the patient. This SPK framework translates disease probabilities into quantitative statements about the ratio of benefits to costs. For a clinician already using the BPP framework, our approach is essentially free: simply invert an odds of disease.

The SPK framework does not replace more advanced decision support models such as Parmigiani [4], Kornak and Lu [5], or Skaltsa et al. [6]. Those methods address more complex decision tasks where either the patient state or the potential action is not binary. The SPK framework is meant to fill a need unmet by those tools: a flexible framework for quantitative decision making that matches the simplicity and ease of use of the BPP framework. In this way, the SPK framework fills much of the same gap as the PK framework [8] yet is easier to use in that it does not require pre-specification and quantification of relevant costs and benefits in order to use.

Beyond clinical practice, we expect that the SPK Framework will be useful in medical education. Much as the BPP framework provides a formal language for interpreting the value of a medical test, the SPK framework provides a formal language for medical decision-making. Educators can use this language to discuss the intricacies of medical decision making and to explain their own decisions to students. Moreover, this framework will clarify the distinction between quantifying the state of a patient and quantifying costs associated with over- and under-treating.

Finally, we proposed a new interpretation of the results of Morgan et al. [10]. Beyond the factors already suggested, we argue that physicians may conflate the probability of disease with the costs of over- and under-treating, which would appear as over-estimation of pre- and post-test probabilities. While we believe this topic requires further study, given the present evidence, we suggest that future studies looking to elicit clinician estimated pre-test or post-test probabilities also survey attitudes towards costs and actions (see Heckerling et al. [21] and Baghdadi et al. [22] for practical examples). Notably, this recommendation has been made elsewhere albeit due to different concerns [19].

There are countless avenues for future study of the SPK framework; we highlight three. First, many papers support the BPP framework by providing quantitative estimate for likelihood ratios of different clinical signs and tests (e.g., Coburn et al. [23]). In fact, these papers can be useful in estimating decision boundary for the SPK framework. Future studies can similarly support both the PK and SPK framework by providing clear statements and quantification of cost-ratios. Beyond this, we note that more objective measures such as hazard ratios or odds ratios represent forms of cost-ratios that can be used in the SPK framework. Second, we have only demonstrated a very restricted set of tools for evaluating cost-ratios. A wide literature on eliciting decision makers preferences could be applied to quantify clinicians’ and patients’ attitudes towards costs [24, 25]. Finally, we have not addressed potential conflicts between clinicians’ and patients’ attitudes towards cost though such conflicts exist. We imagine that future studies may find the SPK framework useful in mitigating these conflicts as it may catalyze discussions about costs.

## Data Availability

No data sets were generated in the present work.

## Additional Files

Supplementary materials and information can be found in the ‘Additional File 1’ pdf.

## Declarations

### Ethics approval and consent to participate

Not applicable.

### Consent for publication

Not applicable.

### Availability of data and materials

Data sharing is not applicable to this article as no datasets were generated or analysed during the current study.

### Authors’ contributions

MPN, JDS, and FM conceptualized the SPK framework and its supporting mathematical details. JDS, CS, and JSS developed the arguments for bacteriuria and breast cancer case studies. MS and CP suggested the connection to Morgan et al. which was further developed by MPN and JDS. MPN and FM found and analyzed the necessary data to support the case studies and implemented any necessary software code. MPN and JDS prepared the original draft. All authors read and approved the final manuscript.

