## Supplemental Materials for "From Pre-test and Post-test Probabilities to Medical Decision Making"

#### 1 Further Mathematical Details of SPK framework

The decision boundary for the SPK framework comes from equating the expected costs of action with the expected costs of inaction, i.e.:

$$\text{Expected Cost(action)} = \text{Expected Cost(inaction)} \tag{1}$$

$$pc_{\text{aa}} + (1-p)c_{\text{ia}} = (1-p)c_{\text{ai}} + pc_{\text{ii}}. \tag{2}$$

Let $r^* = \frac{1-p}{p}$ and $r = \frac{c_{\text{ii}} - c_{\text{aa}}}{c_{\text{ia}} - c_{\text{ai}}}$. Since $r$ can be negative, there are four separate cases to consider when deriving the boundary. To the best of our knowledge, this subtly was not discussed in the original PK framework.

**Case 1:** If $c_{\text{ii}} - c_{\text{aa}} > 0$ and $c_{\text{ia}} - c_{\text{ai}} > 0$, it is optimal to act if $r > r^*$. We will act if

$$pc_{\text{aa}} + (1-p)c_{\text{ia}} < (1-p)c_{\text{ai}} + pc_{\text{ii}} \tag{3}$$

Since neither the cost of over-action or under-action is negative, rearrangement yields:

$$(1-p)(c_{\text{ia}}-c_{\text{ai}}) < p(c_{\text{ii}}-c_{\text{aa}}) \tag{4}$$

$$r^* < r. \tag{5}$$

**Case 2:** If $c_{\text{ii}}-c_{\text{aa}} < 0$ and $c_{\text{ia}}-c_{\text{ai}} > 0$, inaction is always optimal. The constraint implies $c_{\text{aa}} > c_{\text{ii}}$ and $c_{\text{ia}} > c_{\text{ai}}$. Thus, both costs associated with action are larger than the costs associated with inaction, so inaction is always optimal.

**Case 3:** If $c_{\text{ii}}-c_{\text{aa}} > 0$ and $c_{\text{ia}}-c_{\text{ai}} < 0$, action is always optimal. The constraint implies $c_{\text{ii}} > c_{\text{aa}}$ and $c_{\text{ai}} > c_{\text{ia}}$. Thus, both costs associated with inaction are larger than the costs associated with action, so action is always optimal.

**Case 4:** If $c_{\text{ii}}-c_{\text{aa}} < 0$ and $c_{\text{ia}}-c_{\text{ai}} < 0$, it is optimal to act if $r < r^*$. Using a similar argument as in Case 1, we will act if:

$$pc_{\text{aa}} + (1-p)c_{\text{ia}} < (1-p)c_{\text{ai}} + pc_{\text{ii}} \tag{6}$$

However, since the cost of over-action is negative, the sign on the equality flips when rearranging:

$$(1-p)(c_{\text{ia}}-c_{\text{ai}}) < p(c_{\text{ii}}-c_{\text{aa}}) \tag{7}$$

$$r^* > r. \tag{8}$$

#### 2 Breast Cancer Case Study

This case study concerns genetic testing for breast cancer. We demonstrate how to use a cost function based on one element (e.g., mortality) as a benchmark to guide decisions. In Section 2.1, we present the entire case study, and Section 2.2 comprises all supporting mathematical details.

#### 2.1 Prophylactic Mastectomy for a BRCA Carrier

Breast cancer is responsible for over 464,000 deaths per year globally [1]. Genetically, BRCA1 and BRCA2 gene mutations are responsible for 80% of all hereditary breast cancers and 5 to 6% of all cases of breast cancer [2]. Carriers of BRCA1 and BRCA2 have a 40 to 80% lifetime risk of breast cancer [3], much higher than the 8 to 10% risk for the general population [4]. Because of that risk, such patients may consider prophylactic bilateral mastectomy [5]. For many patients, this tremendously difficult decision problem involves a myriad of objective and subjective factors, including mortality, morbidity, psycho-social, and financial costs such as procedural costs and loss of income.

Consider an otherwise healthy 30 year old mother of two with an extensive family history of breast cancer (see Table 1). Based on her genetic test results and her family history, she has an approximately 95.9% post-test probability of being a BRCA1 carrier (post-test odds of 23.8; see Section 2.2.1 for further details). Although the BRCA1 genetic test is highly sensitive and specific, we assume some diagnostic uncertainty remains. Based on prior research, objective measures can capture this patient's mortality risks. In Section 2.2.2, we use prior work to estimate that, in this patient, the expected Years of Life Lost (YLL) for under-treating is 4.40 years and 0.67 years for over-treating. Thus, the mortality-based cost-ratio is $r_{\text{mortality}} \approx 4.40/0.67 = 6.56$ under the assumption that the costs associated with accurate action or accurate inaction are comparatively small. Other factors such as psycho-social and monetary costs remain and are not taken into account in $r_{\text{mortality}}$. In this setting, with some quantified costs, we can apply the SPK framework and create a modified decision boundary to account for these quantified costs, focusing the decision on the remaining unquantified costs.

Placing ourselves in the role of the clinician-patient team, we start by calculating the decision boundary: $r^* = 1/23.8 = 0.04$. Thus in order to warrant action, the cost of under-treatment must be greater than 0.04 times the cost of over-treating. With the boundary less than 1, we choose to invert this statement: *in order to warrant in-action, the cost of over-treating must be* $1/r^* = 23.8$ *times greater than the cost of under-treating.* Evaluating this boundary requires considering at least mortality, morbidity, psycho-social, and monetary factors. However, with a quantified cost-ratio for mortality, we can at least simplify the problem and create a modified decision boundary to evaluate by only considering the remaining unquantified factors (e.g., morbidity, psycho-social, and monetary). Working with inverse cost-ratios again, we have $1/r_{\text{mortality}} = 1/6.56 = 0.152$, far less than $1/r^*$. The comparison implies that if mortality was the only important factor, we should clearly act. We can now consider the other three factors and ask whether, taken together, they provide a strong enough argument for inaction to overcome the mortality cost. We have two potential approaches for this calculation: modifying the decision boundary, or modifying the mortality cost-ratio.

We modify the decision boundary and create a new boundary

$$r^*_{\text{modified}} = \frac{1/r^*}{1/r_{\text{mortality}}} \approx 157.$$

This boundary means: *excluding mortality costs, morbidity, psycho-social, and monetary costs must imply that over-treating is at least 157 times more costly than under-treating to warrant inaction.* Again, mental tricks may help evaluate this modified boundary. As in the prior case-study, this boundary can be phrased in monetary terms: *excluding the mortality cost, if under-treating a patient costs $1000, would you pay at least $157,000 to avoid over-treating?*

Alternatively, we investigate how much we need to modify the mortality cost-ratio to warrant treatment with our original boundary. We can formulate this calculation as a math problem:

$$\frac{0.67 \text{ YLL due to over-treating } + x}{4.40 \text{ YLL due to under-treating} + y} = 23.8.$$

Solving for $x$ gives $x = 104.05 + 23.8y$, which means: *if no other costs for under-treating are present (i.e., if $y = 0$), morbidity, psycho-social, and monetary factors must be equivalent to an additional 104.05 years of life lost due to over-treatment to warrant inaction.* For each additional year of life lost due to under-treating, other factors must contribute an additional 23.8 years of life lost to over-treatment to warrant inaction.

As in the prior bacteriuria case-study, these are just two out of an infinite number of possible ways to present and evaluate this decision boundary. In fact, we expect that some readers will object to our second presentation: as most humans will not live 100 years, considering costs as 104 years of life lost may distort the problem. As in the prior case study, this case-study merely illustrates how extended BPP framework can be a quantitative framework for medical decision-making and highlights ways to use it. Moreover, this case-study illustrates how to incorporate quantified measures of costs with the SPK framework, simplifying the decision problem to the remaining unquantified factors.

#### 2.2 Supporting Mathematical Details

##### 2.2.1 Calculating Post-Test Probability

Highly sensitive and specific tests exist for testing for BRCA. In the case of BRCA1, these tests have a reported average sensitivity of 97.1% (95%-CI: 95.2% - 98.5%) and specificity of 100% (95%-CI: 96% - 100%) [6]. To be conservative, we assume the midpoint of each range in our calculations, resulting in a sensitivity of 96.85% and a specificity of 98%. These percentages result in a positive likelihood ratio of 48.4 ($LR_+$) and a negative likelihood ratio of 0.03 ($LR_-$). Although some tests have no reported false positives (i.e., a specificity of 100%), we do not assume which test was used and, therefore, use a more conservative estimate.

We considered an otherwise healthy 30 year old woman with an extensive familial history of breast cancer (see Table 1) and no family history of ovarian cancer. Every incidence of breast cancer involved a single breast, and each family member was white and not an Ashkenazi Jew. Estimated pre-test probabilities for our case study were computed using the `BayesMendel` package in `R` [7].

Based on her family history, `BayesMendel` returns a 33% pre-test probability of being a BRCA1 carrier (pre-test odds: 0.493). This probability, along with the positive LR above, results in a post-test probability that she is a BRCA1 carrier of 95.9% (post-test odds: 23.839).

| Relationship | BC | Age at Diagnosis | Age at Death | Current Age |
|---|---|---|---|---|
| Sister | Yes | 43 | — | 44 |
| Mother | Yes | 41 | 55 | — |
| Father | No | . | 87 | — |
| Grandmother | Yes | 41 | 45 | — |

**Table 1** The example patient's family history of breast cancer.

##### 2.2.2 Estimating Years of Life Lost

We used a mortality-based metric, the expected years of life lost, in our breast cancer example. We computed it directly from available life expectancy tables in the diseased and non-diseased population.

Consider a hypothetical, currently healthy patient at age $a$. This patient may or may not be diagnosed with breast cancer and may or may not die of breast cancer if diagnosed. Let $t$ denote the age at diagnosis and $s$ denote the age at death if diagnosed. Following the basic properties of expectation, we write the expected years of life lost as the expected difference in the life expectancy for the average person at age $a$ ($LE_a$) and the age at death for those that are diagnosed:

$$E[YLL_i] = E[(LE_a - s)] \\ = \int_{s=a}^{100} (LE_a - s) \left( \int_a^{80} p(s|t)p(t)dt \right) ds. \tag{9}$$

This formulation allows us to incorporate uncertainty in the age at diagnosis into the calculation (as seen in the parenthesis of Equation 9). For BRCA carriers, this metric can be readily calculated from pre-existing data, including the general population life expectancy tables [8], age at diagnosis data for BRCA carriers [9], and death rates following years since diagnosis for breast cancer [10].

We can calculate an analogous measure for expected years of life lost if treatment is pursued but the person is not diseased (over-treatment). In our case study, this is the expected years of life lost if a person pursues a prophylactic mastectomy but does not carry a BRCA mutation, i.e. the death rate for the preventative surgery. We obtained data for this calculation from El-Tamer et al. [11].

We make several assumptions: diagnoses, if they occur, happen before the age of 80 (as noted on the bounds of integration in the parenthesis in Equation 9); the maximum lifespan is 100 years old (as noted on the bounds of integration outside of the parenthesis); and survival for more than 10 years after diagnosis constitutes a "cure" from breast cancer.

We used Monte Carlo simulation to estimate these measures. The general procedure to estimate years of life lost to breast cancer in a BRCA carrier who forgoes an prophylactic mastectomy is in Algorithm 1. For each estimate, we used 1,000 samples.

**Algorithm 1** Estimating years of life lost due to breast cancer for BRCA carriers who do not seek preventative treatment.

```
1: Data: Current age of patient: a
2: Number of Monte Carlo samples: n_samps
3: Result: Expected years of life lost: E(YLL)
4: for i = 1, ..., n_samps do
5:     Draw an indicator if the patient will develop breast cancer, i.e., Y ~ Bern(π)
6:     if Y == 1 then
7:         Draw the age at diagnosis, t_i ~ f_diagnosis
8:         Draw the additional amount of years lived past diagnosis, δ_i ~ f_survival since diagnosis
9:         if δ_i < 10 then
10:            Set age at death s_i = t_i + δ_i
11:        else
12:            Declare a "cure" and draw the age at death from the general population, i.e. s_i ~ f_general pop
13:        end if
14:    else
15:        Draw the age at death from the general population, s_i ~ f_general pop
16:    end if
17:    Draw an age at death for the general population, LE_i ~ f_general pop
18:    Calculate the expected years of life lost, YLL_i = LE_i − s_i
19: end for
20: Return mean(YLL)
```

### 3 Additional Details for Re-Analyzing Morgan et al.

#### 3.1 Deriving Implied Cost Ratio

We show that the implied cost ratio in our re-analysis of Morgan et al. [12] is $r = o_{\text{act}}/o_{\text{disease}}$. This result assumes that the odds that a physician will act is proportional to their perception of the expected costs:

$$o_{\text{act}} = \frac{\text{Expected Cost of Not Treating}}{\text{Expected Cost of Treating}}.$$

Combining the above ratio for $r$ with the assumption above, we obtain

$$r = o_{\text{act}}/o_{\text{disease}} = \frac{\text{Expected Cost of Not Treating}}{\text{Expected Cost of Treating}} \times \frac{1}{o_{\text{disease}}},$$

which, using the relationship between probability and odds, implies:

$$r = o_{\text{act}}/o_{\text{disease}} = \frac{\text{Expected Cost of Not Treating}}{\text{Expected Cost of Treating}} \times \frac{1 - p_{\text{disease}}}{p_{\text{disease}}}.$$

Recall that the expected cost of not treating is $(1 - p_{\text{disease}})c_{ai} + p_{\text{disease}}c_{ii}$, and the expected cost of treating is $p_{\text{disease}}c_{aa} + (1 - p_{\text{disease}})c_{ia}$. Under the assumption that the cost of the correct decision is minimal (i.e., $c_{aa} \approx 0$ and $c_{ai} \approx 0$), this is exactly the equation presented in the main text as it cancels with the second term on the right-hand side of the equation above:

$$r = \frac{p_{\text{disease}}c_{ii}}{(1 - p_{\text{disease}})c_{ia}} \times \frac{1 - p_{\text{disease}}}{p_{\text{disease}}} = \frac{c_{ii}}{c_{ia}} = \frac{c_{\text{under-action}}}{c_{\text{over-action}}}.$$

#### 3.2 Exclusion of the UTI Survey Results

In our re-analysis of Morgan et al. [12], we disregard their UTI case study, due to both survey design and corresponding cited literature. The presented clinical scenario was as follows:

*"Mr. Williams, a 65-year-old man, comes to the office for follow up of his osteoarthritis. He has noted foul-smelling urine and no pain or difficulty with urination. A urine dipstick shows trace blood. He has no particular preference for testing and wants your advice."*

In this scenario, the patient presented with both blood in the urine and foul-smelling urine, yet the researchers assumed asymptomatic bacteriuria. The predictive power of these symptoms are often noted as controversial or misleading in the medical literature (e.g., Midthun et al. [13], Jump et al. [14]), yet around half of family practice physicians consider them when considering antibiotic therapy for UTIs [15]. Indirectly, the discussion of Morgan et al. notes this as the differences observed in this example "may reflect the evolution of the definition of asymptomatic bacteriuria as a separate entity from UTI" [12]. Thus, we exclude the asymptomatic bacteriuria example from our reanalysis of Morgan et al. [12].

##### 3.3 More Notes on the Breast Cancer Survey Cost-Ratio

Though the clinical scenario is somewhat vague with a positive mammogram but no mention of the size or differential diagnosis, the next logical action is to pursue a confirmatory diagnosis with a core needle biopsy. We are unaware of studies studying the mortality risk associated with not doing a biopsy in a patient with cancer (e.g., under-treating); we assume that if this patient had cancer but did not receive a biopsy, she would also not undergo surgery. Prior work studying mortality risk in patients who have breast cancer but refuse surgery (under-treating) suggest that the 5 year mortality risk is approximately 30% [16]. In contrast, over-treating consists of mortality risk associated with a core-needle biopsy combined with mortality risk of developing a new breast-cancer after a false-positive mammogram. Since the mortality risk associated with a core-needle biopsy is minuscule, we focus on 5-year mortality risk from breast-cancer in patients after receiving a false-positive mammogram. For women who do not have breast cancer, less than 1% are expected to develop breast-cancer within the next 5-years [17]. Thus, we have an absolute upper bound of a 1% 5-year mortality rate associated with developing breast cancer in the setting of over-treating. Thus, the cost-ratio for this patient should be at least $CR = 0.30/0.01 = 30$, which is above the implied cost-ratio of 17. In short, these results do not disprove our hypothesis that bluntness of the survey tool contributed to the physicians' elevated probability estimates.

##### 3.4 Calculating Cost Ratio using Heckerling et al.

In Heckerling et al. [18], 52 physicians reviewed the charts of their own patients who had presented to an emergency department with fever or respiratory complaints. They had considered whether to order chest x-rays for diagnosing pneumonia. They asked each physician to assign a cost to correctly treating, under-treating, and over-treating in each scenario, by ranking each from -50 (described as the worst possible action) to 50 (described as the best possible action). On average, attending physicians reported the cost of under-treating as -45, and the cost of over-treating as 5.

We calculated a cost ratio of under-treating to over-treating, using the data for attending physicians. We re-scaled the data points between 0 and 100, applied the logit transformation, and calculated the cost ratio by dividing the absolute logit-transformed cost of under-treating by the absolute logit-transformed cost of over-treating. We found an estimated cost ratio of 14.7.

While this result is slightly lower than the 15.8 cost-ratio we estimated from Morgan et al. [12], we consider the correspondence (14.7 versus 15.8) remarkable. Overall, this result strongly suggests that future work exploring the cause of physicians' inflated probability estimates should consider that physicians may conflate probabilities of disease with factors affecting decisions to act.

[4] Carbonara N, La Forgia D, Pellegrino R, Ressa C, Tommasi S. A cost decision model supporting treatment strategy selection in BRCA1/2 mutation carriers in

breast cancer. J Pers Med. 2021;11(9):847.

[5] Elezaby M, Lees B, Maturen KE, et al. BRCA mutation carriers: breast and ovarian cancer screening guidelines and imaging considerations. Radiology. 2019;291(3):554–569.

[6] U S CDC.: BRCA and Breast/Ovarian Cancer Analytic Validity. Available from: https://www.cdc.gov/genomics/gtesting/file/print/fbr/bcanaval.pdf.

[7] Parmigiani G, Chen S, Wang W, et al.: BayesMendel: Determining carrier probabilities for cancer susceptibility genes. R package version 2.1-8-1. Available from: https://projects.iq.harvard.edu/bayesmendel.

[8] Social Security.: Actuarial Life Table: A Period Life Table on Mortality. https://www.ssa.gov/oact/STATS/table4c6.html.
